# Willingness to Accept Tradeoffs among Covid-19 Cases, Social-Distancing Restrictions, and Economic Impact: A Nationwide US Study

**DOI:** 10.1101/2020.06.01.20119180

**Authors:** Shelby Reed, Juan Marcos Gonzalez, F. Reed Johnson

## Abstract

We designed a discrete-choice experiment to quantify the extent to which US adults would accept greater risk of infection with SARS-CoV-2 in return for lifting social-distancing restrictions and diminishing the economic impact of the COVID-19 pandemic. 5953 adults recruited from the SurveyHealthcareGlobus internet panel representing all 50 states had 4 distinctly different preference patterns. About 37% were risk minimizers reluctant to accept any increases in risk of contracting the virus. Another group (26%) was primarily concerned about time required for economic recovery, accepting increases in COVID-19 risk levels up to 16% to shorten recovery from 3 to 2 years. The remaining two groups diverged on the relative importance of reopening nonessential businesses. The larger group (26%) strongly preferred delaying reopening while the smaller group (13%) would accept COVID-19 risks well beyond 20% to avoid a delay in reopening. Political affiliation, race, household income and employment status were predictive of group membership.

## Introduction

The economic damages to the economy have become increasingly more worrisome under social-distancing restrictions designed to combat the spread of SARS-CoV-2. Despite models predicting increases in COVID-19 infections as restrictions are lifted and risks of local spikes in cases, Governors across the United States are experimenting with phased restarting of economic activity. These decisions require explicit judgments about the relative importance of the risks associated with COVID-19 cases and the economic toll of social-distancing restrictions with downstream effects on non-COVID-19 health outcomes, social instability, poverty, and provision of public services such as education and law enforcement.

While the preferences of a few are clearly exhibited in protests at state capitals while others remain isolated in their homes, decision makers lack information from studies designed to systematically evaluate the public’s views on the tradeoffs between COVID-19 management policies and their economic impact. Public-opinion surveys indicating general support for or opposition to social distancing do not provide information on the value judgements required to evaluate the timing and scale of lifting restrictions. Rigorous quantification of the public’s acceptance of these tradeoffs could provide valuable information to government and public health officials during the initial phase of the pandemic as well as in response to subsequent spikes in infections. Lifting restrictions may not immediately induce risk-averse people to re-engage in economic activities. However, identifying the sizes of groups with distinctive preferences for health versus economic tradeoffs also could help decision makers assess the likely benefits and costs of various policies.

## STUDY DATA AND METHODS

We designed a discrete-choice experiment to determine the extent to which Americans are willing to accept greater spread of SARS-CoV-2 to lift social-distancing restrictions and limit the economic impact of the pandemic. We hypothesized that preferences would differ by sex, age, education, political affiliation, household income, and employment status.

### Survey development

The discrete-choice experiment focused on four factors: COVID-19 risk, the duration of social-distancing restrictions, and the depth and duration of negative economic impacts. COVID-19 risk was described as the percentage of the US population infected with the virus through the end of 2020 accounting for all cases, confirmed and unconfirmed. Levels ranged from 2% to 20%, based on an estimated 1.4 million diagnosed cases on May 14, 2020 in the US, increased by factors of approximately 5 to 50 to account for undiagnosed cases and potential increases through 2020.^1^

We asked respondents to consider the duration of restrictions on nonessential businesses such as hair salons, fitness clubs and retail stores. For this factor, respondents were told when restrictions for these businesses would be lifted with levels ranging from “now” (May 2020) to “October” representing a duration of five additional months. The depth of the economic toll was portrayed as an increase in the percentage of US households falling below the poverty threshold, from 13%, the national average in 2018.^2^ Three levels included increases to 16%, 20% and 25%. The fourth factor representing the duration of economic impact was described as the number of years before the economy would recover to “pre-COVID-19 levels”, with 2-year, 3-year and 5-year levels.^3^ An example choice question is provided in the supplemental materials.

To provide context about the relative importance of different types of social-distancing restrictions, the survey also included a ranking exercise in which respondents were asked to rank-order the importance lifting six groups of restrictions: 1) reopening nonessential businesses; 2) allowing dine-in meals in restaurants; 3) reopening schools and colleges; 4) allowing sporting events to resume; 5) allowing religious ceremonies to resume; and 6) reopening parks and museums. Additional survey items were included to collect sociodemographic characteristics, 3-digit ZIP code, health conditions and other information possibly related with respondents’ preferences.

We used Lighthouse Studio Version 9.8 (Sawtooth Software; Orem, UT) to program the web-based survey. Factor levels shown across choice questions were governed by an orthogonal experimental design with 300 versions of 10 pairs of hypothetical scenarios. Each respondent was assigned to one of the 300 versions and answered 10 choice questions. To field the survey, we collaborated with SurveyHealthcareGLOBUS, a healthcare market-research firm. SurveyHealthGLOBUS sent emails to adults across the U.S. inviting them to complete the survey, with oversampling in New York, California, Florida, Texas, and North Carolina.

The study protocol was reviewed and determined to be exempt by the Duke Health Institutional Review Board (Pro00105431). There was no external funding for the study.

### Analysis

Given the wide range of views expressed in the lay press and social media outlets, our statistical modeling approach focused on exploring and characterizing heterogeneity in preferences across respondents. We used conditional-logit latent-class analysis to identify groups of respondents with similar choice patterns. In preliminary models, categorical models indicated that COVID-19 risk levels were linearly associated with the log-odds of chosen alternatives. Subsequently the COVID-19 regression parameter was modeled as a linear, continuous covariate. The relative importance of other factors are reported as the increases in COVID-19 risk levels that were perceived to the have the same importance as changes in duration of nonessential business closures, increases in the percentage of households below the poverty line, and longer durations of a COVID-related economic downturn. To test whether sex, age, education, political affiliation, household income, and employment status were associated with membership in different preference groups, we added respondent-level characteristics to the latent-class models as covariates. We used Stata/SE 16.1 (StataCorp LLC; College Station) and LatentGOLD 5.1 (Statistical Innovations Inc.; Belmont, MA) software for analysis.

## RESULTS

5953 respondents completed the survey between May 8 and May 20, 2020. Respondents were generally representative of the adult US population. **Table 1** provides descriptive statistics for our study cohort in relation to the US adult population. Our sample matched the national age distribution, but was more likely to be female and white, and generally had more formal education than the general population. However, our study cohort had large numbers of respondents of varying ages and income levels with wide geographic dispersion representing all 50 states. Approximately 4 in 10 identified as Democrat, 3 in 10 as Republican and nearly 3 in 10 as independent political affiliation.

**Table 1.** Characteristics for US Adult Population and Survey Respondents

|  | **US Adult**<br>**Population** | **Survey**<br>**respondents**<br>**N=5,953** |
| --- | --- | --- |
| Female | 52%<sup>a</sup> | 67% |
| Age, mean | 47 years<sup>a</sup> | 48 years |
| 18-44 | 46% | 45% |
| 45-64 | 33% | 35% |
| 65 years and older | 21% | 20% |
| Race |  |  |
| White | 63%<sup>b</sup> | 77% |
| African American | 12% | 8% |
| Hispanic or Latino | 16% | 6% |
| Asian | 6% | 5% |
| Other<sup>c</sup> | 3% | 3% |
| Education |  |  |
| High school or less | 39%<sup>d</sup> | 20% |
| Some college | 18% | 20% |
| Associate degree | 10% | 17%<sup>e</sup> |
| Bachelor degree | 21% | 26% |
| Graduate degree | 12% | 18% |
| Household income |  |  |
| <\$25,000 | 17%<sup>a</sup> | 18%<sup>g</sup> |
| \$25,000 to \$49,999 | 21% | 25% |
| \$50,000 to \$99,999 | 29% | 35% |
| \$100,000 or more | 32% | 22% |
| Geographic region |  |  |
| Northeast | 18%<sup>a</sup> | 21% |
| Midwest | 21% | 23% |
| South | 38% | 37% |
| West | 24% | 19% |
| Political affiliation |  |  |
| Democrat | 31%<sup>h</sup> | 40%<sup>g</sup> |
| Independent | 36% | 29% |
| Republican | 30% | 32% |

<sup>a</sup> U.S. Census Bureau, Current Population Survey, Annual Social and Economic Supplement, 2018
<sup>b</sup> Adult population by race in 2018. From National KIDS COUNT. Population Division, U.S. Census Bureau. <sup>c</sup> includes American Indian, Alaskan native, native Hawaiian, other Pacific Islander, and mixed race<sup>f</sup>
<sup>d</sup> U.S. Census Bureau, Current Population Survey, Annual Social and Economic Supplement, 2019
https://datacenter.kidscount.org/data/tables/6539-adult-population-by-race#detailed/1/any/false/37/68,69,67,12,70,66,71,2800/13517,13518
<sup>e</sup> Associate's degree and technical college combined
<sup>f</sup> includes furloughed (162), laid off (139), student (192), other (141), military (9), homemaker (476), and "prefer not to say" (34)
<sup>g</sup> excludes from the denominator 5% of respondents who 'did not know' or 'prefer not to say'.
<sup>h</sup> Gallup. Party Affiliation. April 14-28, 2020. Available at: https://news.gallup.com/poll/15370/party-affiliation.aspx

Respondents answered a simple ranking question on the relative importance of reopening various activities. On average, respondents considered reopening nonessential businesses to be the most important (mean ranking 2.3, with 1 being most important) with 37% ranking it as most important policy. Reopening schools and colleges was second most important (mean, 3.2), followed by allowing dine-in meals in restaurants (mean, 3.5), reopening parks and museums (mean, 3.5), allowing religious services to resume (mean, 3.7) and allowing sporting events to resume (mean, 4.8). All pairwise differences were statistically significant (p<0.0001) with the exception of restaurants versus parks/museums.

Model-fit statistics indicated that a four-class model provided good data fit with relatively large segments of the sample distributed across all four classes. **Figure 1** shows variations in the relative importance of the four factors used to portray alternative scenarios. Class 1 represented approximately 36% of respondents who predominantly selected scenarios with the lower cumulative incidence of COVID-19. For ease of exposition, we refer to this group as “risk-minimizers”. Class 2 and Class 3 were similarly sized, with approximately 26% and 25% of the sample in these groups, respectively. Class 2 was the only group with positive preferences for delaying the reopening of non-essential businesses until October, independent of COVID-19 risks, and they gave more weight to the poverty factor. Respondents in class 3 prioritized scenarios depicting faster economic recovery; they placed little importance on when nonessential businesses would be reopened. For this class, reducing the time required for economic recovery from 5 years to 2 years was about twice as important as reducing the cumulative risk of COVID-19 cases through 2020 increasing from 2% to 20%. For ease of exposition, we refer to class 2 as “waiters” and class 3 as “pro-business”. Class 4 represented about 13% of respondents who strongly prefer reopening nonessential businesses now. Hence, we refer to this class “openers”.

**Figure 1.**
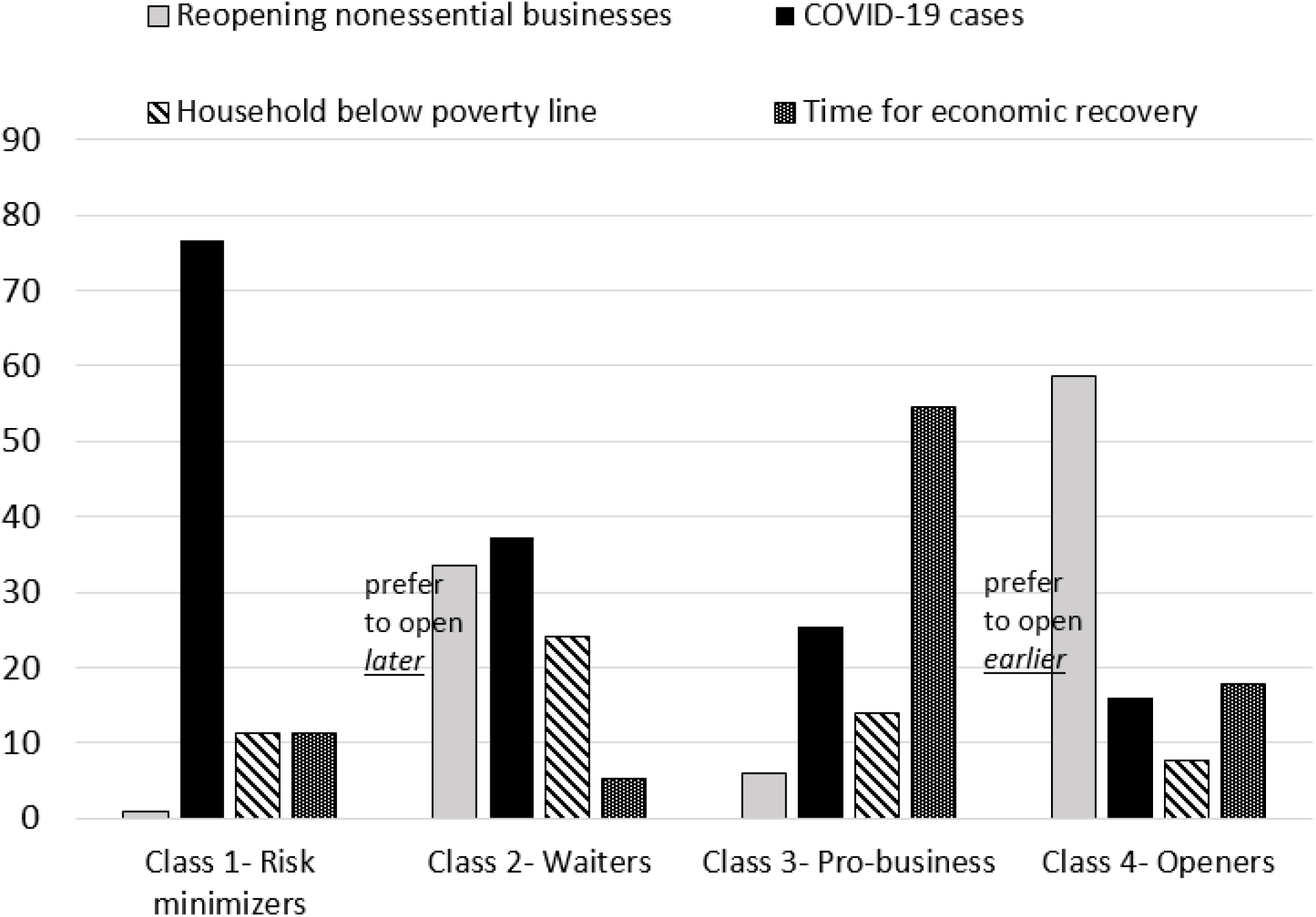
Relative importance of discrete-choice experiment factors

Dividing preference weights by the COVID-19 slope, indicating the marginal utility of a 1-percent change in risk, allows us to report the relative importance of economic gains scaled as the corresponding maximum risk of COVID-19 respondents in each group would accept in return for these gains. This maximum-acceptable risk metric is directly analogous to the way willingness to pay is calculated in choice-experiment studies that include a cost factor.^5^ The maximum-acceptable risks for the COVID-19 risk-minimizer group are very small for improvements on the other three factors because individuals with these preferences are not willing to accept COVID-19 risk regardless of when non-essential businesses are reopened, how many fewer families fall below the poverty line, or how much faster the economy recovers (**Figure 2**). Since the class referred to as ‘waiters’ preferred delaying reopening nonessential businesses, there also is no level of COVID-19 risk they would find acceptable for lifting this restriction earlier. People with these preferences, however, were willing to accept a 5.9 percentage-point increase in risk of COVID-19 to avoid a 4 percentage-point increase in households below the poverty level. This finding was similar for people with pro-business and opener preferences, who willing to accept a 4 to 5 percentage-point increase in COVID-19 risk to avoid a 4% percent increase in households in poverty.

**Figure 2.**
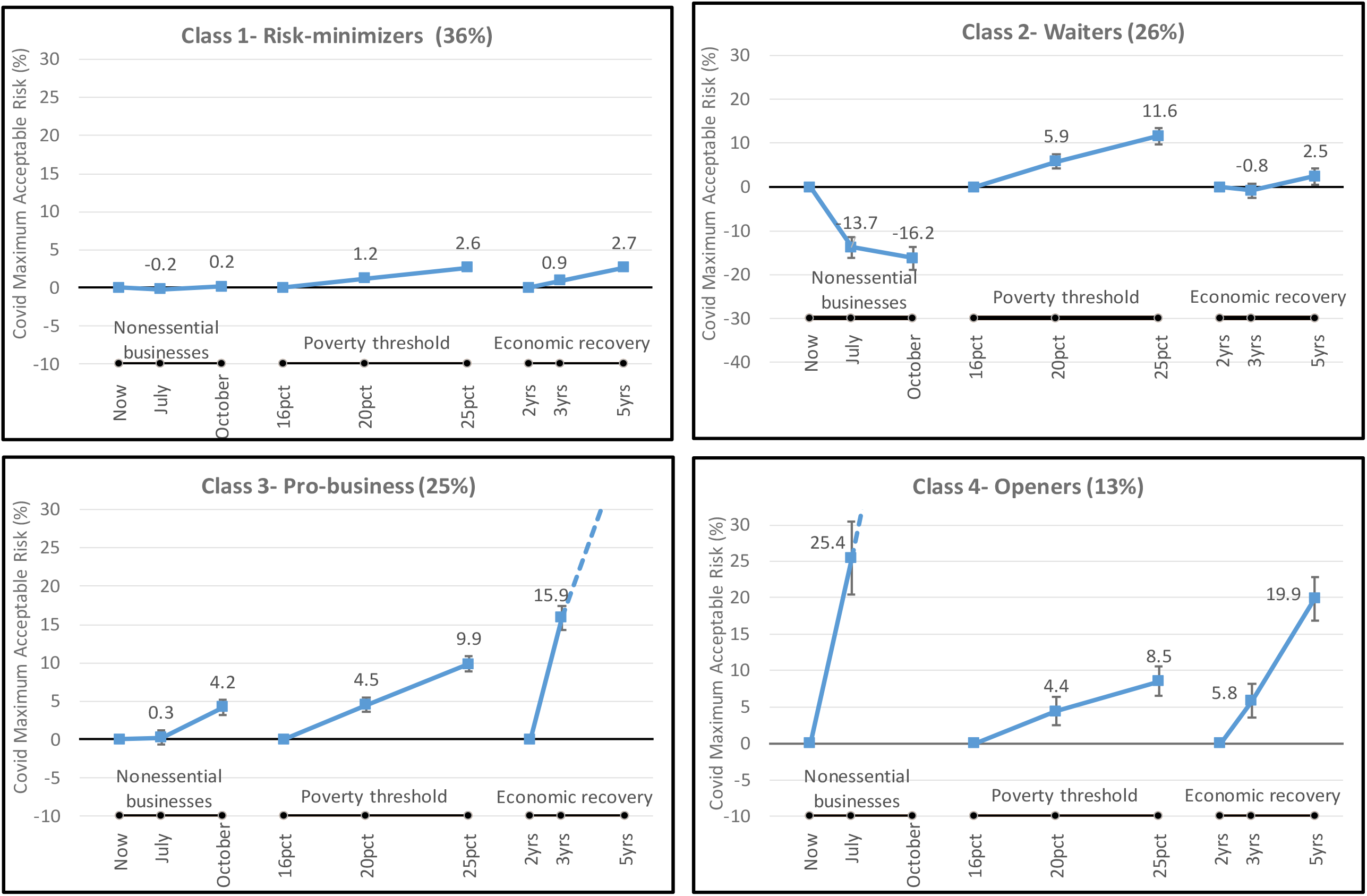
Maximum-acceptable cumulative risk of COVID-19 by preference group

These two groups, however, were willing to accept higher levels of COVID-19 risk to hasten economic recovery. The people with pro-business preferences would accept a 15.9% increase in the risk of COVID to reduce the time for economic recovery by at least 1 year (from 3 years to 2 years), and the people with openers preferences would accept a 5.8% increased risk of COVID-19 for the same gain. The factor that clearly distinguished these two groups was the importance they placed on reopening nonessential businesses. The people with pro-business preferences gave little regard to this factor, while the openers were willing to accept COVID-19 risks above the 20% upper bound of the range shown in the choice questions. By extrapolating, individuals with these preferences would accept a 25% cumulative risk of COVID-19 to reopen essential businesses now rather than July and even greater risk to avoid waiting until October.

**Table 2** summarizes characteristics of respondents more and less likely to belong to each latent class. The respondent characteristic most strongly associated with class membership was political affiliation (Wald score, 217; *p*<0.0001). Respondents with Democratic and Republican political affiliation both were more likely to be among the risk-minimizers group, while those with independent political affiliation were significantly less likely to belong in this group.

Independents, however, were positively associated with being in the pro-business and people with openers preferences, and they were negatively associated with being in the waiters group. Higher income was positively associated with the people with pro-business preferences and negatively associated with the waiters group. Lower income earners were over-represented in the waiters class while individuals with salaries ranging from $25,000 to $100,000 were more likely to be represented in the openers class. Salaried individuals were less likely to be risk minimizers and openers, but more likely to be in the waiters class. Non-whites were strongly associated with membership in the waiters class while negatively associated with membership in the pro-business and openers classes. Sex and level of education were not independently associated with class membership.

**Table 2.** Respondent characteristics associated with each preference group^a^

| Less likely | More likely |
| --- | --- |
| **Class 1- Risk-minimizers** |  |
| Independent political affiliation**<br>Salaried employment* | Democrat**<br>Other employment status*<br>Retired or disabled*<br>Republican* |
| **Class 2- Waiters** |  |
| Independent political affiliation**<br>Income > \$100,000 per year**<br>Retired or disabled*<br>Republican* | Nonwhite**<br>Democrat**<br>Salaried employment**<br>Income < \$25,000 per year** |
| **Class 3- Pro-business** |  |
| Democrat*<br>Nonwhite* | Independent political affiliation*<br>Income > \$100,000 per year* |
| **Class 4- Openers** |  |
| Democrat**<br>Nonwhite**<br>Income < \$25,000 per year*<br>Salaried employment* | Independent political affiliation**<br>Income \$25,000 - \$100,000 per year*<br>Self-employed* |

<sup>a</sup> Characteristics in each class listed in descending order of magnitude. \*p<0.01; \*\*p<0.001

## DISCUSSION

Governors of all 50 states recently have begun to allow retail shops, dine-in restaurants, salons and fitness facilities to reopen with safety precautions in place to control the spread of COVID-19. Opinion polls throughout the pandemic have cited broader support of social-distancing measures among Democrats followed by independents and Republicans.^4^ However, when acceptance of social-distancing measures are framed in the context of tradeoffs among COVID-19 risks, longer economic downturns, and more families falling below the poverty line, we found that self-identified Democrats and Republicans actually had more similar tradeoff preferences compared to those identifying as independents. This suggests that differences in relative preferences for policies across members of the democratic and republican parties may have more to do with expectations of the impact that policies will have on COVID cases, and less to do with their judgment about acceptable tradeoffs.

Our study, using a nationally representative sample, provides robust, conceptually sound estimates of Americans’ stated willingness to accept increased risks of COVID-19 through 2020 to hasten economic recovery and limit its impact on low-income populations. The large sample size provided ample power to characterize groups of respondents with similar preference patterns and to estimate precise relative-importance parameters. Nearly four in ten respondents prioritized minimizing the risk of COVID-19, and thus were unwilling to accept any level of risk to reopen nonessential businesses or blunt the economic impact of the pandemic. About 44% of Democrats and 40% of Republicans in our sample were predicted to be in this group compared to 27% of independents.

Overall, about one in four respondents chose scenarios consistent with faster economic recovery, with class memberships of 20% of Democrats, 24% of Republicans, and 30% of independents in our sample. This group considered the differences in risk levels shown for COVID-19 to be about twice as important as differences between 2 and 5 years needed for the economy to recover.

The “waiters” class placed relatively little importance on minimizing the risk of COVID-19 but strongly preferring to delay reopening nonessential businesses until the fall initially appeared paradoxical. However, this class was robust across model specifications, was relatively large, and continued to yield multiple significant covariates predictive of class membership. Low-income, non-white, Democrats, and salaried employment covariates were positively associated with class membership. Salaried individuals presumably would be less likely to suffer financially if business openings were delayed. It also is reasonable to assume that low-income individuals would have little means to spend on dine-in eating, salons and fitness centers; thus, having less interest in opening nonessential businesses. This sentiment may also be reflected in their relative disregard for the economic-recovery factor. Since they already are at or below the poverty line, economic recovery could hold little promise for them. The preference profile of this group may also be indicative of individuals hoping for a near-term effective treatment or vaccine. If such a treatment were to become available, then the projected number of COVID-19 cases would not materialize and the economy would rebound as a result.

The final group represented individuals who expressed a strong desire to open the doors to nonessential businesses immediately. Although this implies acceptance of high levels of COVID-19 risk, these individuals may have assumed that they could personally protect themselves to reduce their personal risk of COVID-19 thereby disregarding the actual risk levels shown. Interestingly, this group was not swayed by the impact on poor families, as the poverty factor was the least important. One in five independents (20%) in our sample was likely to belong to this class, compared to 12% of Republicans and 6% of Democrats.

### Limitations

The hypothetical nature of our study is a limitation, as it is with all stated-preference studies. However, these studies can be designed to provide insights on issues that shape public sentiment and behaviors of individuals that are isolated from their everyday reality. For instance, an individual may have strong preferences for lifting social-distancing restrictions and would be willing to assume substantial risks of contracting COVID-19. However, they still may not be willing to violate states’ stay-at-home orders. Others may give priority to restarting the economy and accept high rates of COVID-19 infection because they personally will stay isolated to protect themselves from exposure to the virus.

Although not a limitation of stated-preference studies, respondents could have considered external information when responding to the choice questions in the survey given the nearly continuous coverage of the COVID-19 pandemic across lay and scientific media outlets. Some respondents could have considered relationships between factor levels shown in alternative profiles. For instance, some respondents may have assumed that the percentage of families dipping below the poverty threshold would be lower than shown or placed greater responsibility for economic prosperity among individuals when profiles depicted scenarios with faster economic recovery. However, we checked for statistical interactions between COVID-19 risk and social distancing policies and between time for economic recovery and poverty threshold variables, but these terms were not statistically significant.

### Policy implications

The number of simulation models forecasting future COVID-19 cases is proliferating as the evidence base develops on diagnostic testing, transmission efficiency, health care capacity, and the effectiveness of alternative social-distancing restrictions.^6^ These models do not, however, incorporate the economic costs associated with the pandemic nor the public’s willingness to accept greater infection risks to limit the economic fallout. We hope that the results of our study can support government and public health officials who must make the difficult decisions about when to tighten and when to loosen social-distancing restrictions. Our findings reveal useful information about segments of the population that will be more and less supportive of various policies, findings that do not necessarily track with information reported on social media and traditional media outlets.

Although highly publicized protests and opinion polls have signaled political affiliation as a strong determinant of attitudes toward social-distancing measures, when evaluating explicit tradeoffs among controlling the pandemic, social-distancing restrictions, and economic recovery, we find that Democrats and Republicans are more similar than those who identify as independent.

## Data Availability

Contact the corresponding author for access to the data.

## Notes

### Competing Interest Statement

The authors have declared no competing interest.

### Funding Statement

There was no external funding. SurveyHealthcareGlobus provided in-kind support in the form of access to their internet panel for administering the survey.

### Author Declarations

Duke Health Institutional Review Board (Pro00105431)

